# Nitric oxide gas inhalation to prevent COVID-2019 in healthcare providers

**DOI:** 10.1101/2020.04.05.20054544

**Authors:** Stefano Gianni, Bijan Safaee Fakhr, Caio Cesar Araujo Morais, Raffaele Di Fenza, Grant Larson, Riccardo Pinciroli, Timothy Houle, Ariel Louise Mueller, Andrea Bellavia, Robert Kacmarek, Ryan Carroll, Lorenzo Berra

## Abstract

**Introduction:** In human hosts, SARS-CoV-2 causes a respiratory syndrome (named COVID-19) which can range from a mild involvement of the upper airways to a severe pneumonia with acute respiratory syndrome that requires mechanical ventilation in an intensive care unit (ICU). Hospital-associated transmission is an important route of spreading for the SARS-CoV-2 virus and healthcare providers are at the highest risk. As of February 2020, 1716, Chinese healthcare workers had confirmed SARS-CoV-2 infections and at least 6 died. Unfortunately, there is currently no vaccine or pharmacological prophylaxis to decrease the risk of healthcare providers contracting the infection.

**Methods and analysis:** We will randomize 470 healthcare providers scheduled to work with COVID 19 patients to receive nitric oxide gas administration (NO group, n=235) or no gas administration (control group, n=235). The primary endpoint of this study is the incidence of subjects with COVID-19 disease at 14 days from enrollment. Secondary endpoints are the proportion of healthcare providers who present a positive real time RT-PCR test for SARS-CoV- 2 14 days after enrollment, the proportion of healthcare providers requiring quarantine, and the total number of quarantine days in the two groups.

**Ethics and dissemination:** The trial protocol is under the approval of The Partners Human Research Committee of Massachusetts General Hospital (Boston, USA) and recruitment is expected to start in April 2020. The results of this study will be published in scientific journals and presented at scientific meetings.

## Introduction

The current COVID-19 pandemic started in December 2019 from Wuhan, China and rapidly diffused throughout the Asian continent, to Europe, and more recently to the United States [1]. COVID-19 is predominantly a respiratory infection that spans from a mild involvement of the upper respiratory tract to a severe pneumonia leading to respiratory distress, shock, and death [2].

The disease is commonly transmitted in hospital settings and healthcare providers have the highest risk of being infected. In February 2020, SARS-CoV-2 was confirmed in about 1,700 Chinese healthcare workers, [3] and, on April 4 2020, up to 189 Massachusetts General Hospital employee were infected by SARS-CoV-2. [4] Unfortunately, there is currently no vaccine or pharmacological prophylaxis to decrease the risk of infection in healthcare providers. Thus, a safe and prophylactic therapy to reduce the instance of COVID-19 disease in healthcare workers would be of great benefit to clinical staff and society.

Nitric oxide (NO) is an anti-infective agent at high concentrations. *In-vitro* studies have identified significant activity of NO against pathogens broadly resistant to antibiotic therapy, such as bacteria responsible for hospital acquired pneumonia [5]. In a small-sized clinical trial, nitric oxide gas was shown to be safe and effective against bacterial colonization/superinfections in the context of chronic respiratory diseases [6]. NO gas also exhibited anti-viral activities against SARS-CoV [7] [8]. In an *in-vitro* study, the NO-donor compound S-nitroso-N- acteylpenicillamine increased the survival rate of mammalian cells infected with SARS-CoV [9]. In patients affected by SARS, NO inhalation decreased the need for oxygen therapy, ventilatory support and a faster resolution of chest x-ray abnormalities was observed [10].

Due to the genetic similarities, both Coronaviridae SARS-CoV and SARS-CoV-2 might behave similarly to NO gas exposure. We hypothesize that a high dose of exogenous inhaled NO is a viricidal agent in COVID-19 disease.

A dose of 160 ppm of nitric oxide has been found to be bactericidal, fungicidal, viricidal [11,12], and safe for patients who are adequately managed. High doses of NO administered continuously for long periods of time can generate methemoglobinemia. Miller et al found that intermittent administration of 160 ppm NO for 4 cycles of 30 min have the same antibacterial effect and can be safely administered [13].

We hypothesize that the administration of high doses of nitric oxide will prevent the development of COVID-19 disease in healthcare providers exposed to SARS-CoV-2 positive patients.

## Methods and analysis

### Study setting

This is a single center, randomized (1:1) controlled, parallel-arm clinical trial. Given the absence of a proven prevention strategy for healthcare workers, we propose this protocol to all interested centers with SARS-CoV-2 patients.

### Eligibility Criteria

In-hospital healthcare providers will be recruited. Candidates should be scheduled to work with confirmed SARS-CoV-2 patients for at least 3 days of the week. Exclusion criteria are proven previous SARS-CoV-2 infection and subsequent negative rt-PCR test, pregnancy, known hemoglobinopathies and known anemia. Inclusion and exclusion criteria are summarized in table 1.

**Table 1:** inclusion and exclusion criteria. RT-PCR = real time polymerase chain reaction.

| Inclusion criteria |
| --- |
| Age ≥18 years |
| Scheduled to work with SARS-CoV-2 RT-PCR patients for at least 3 days in a week |
| Exclusion criteria |
| Proven previous SARS-CoV2 infection and subsequent negative RT-PCR test |
| Pregnancy |
| Hemoglobinopathy |
| Anemia |

### Interventions

Eligible subjects will be randomized to receive nitric oxide administration (treatment group) or no gas administration (control group). NO will be delivered in 2 daily sessions (before and after the work shift) for 14 consecutive days. Each session will last 15 minutes, for a total of 30 minutes/day for each subject. See figure 1.

**Figure 1.**
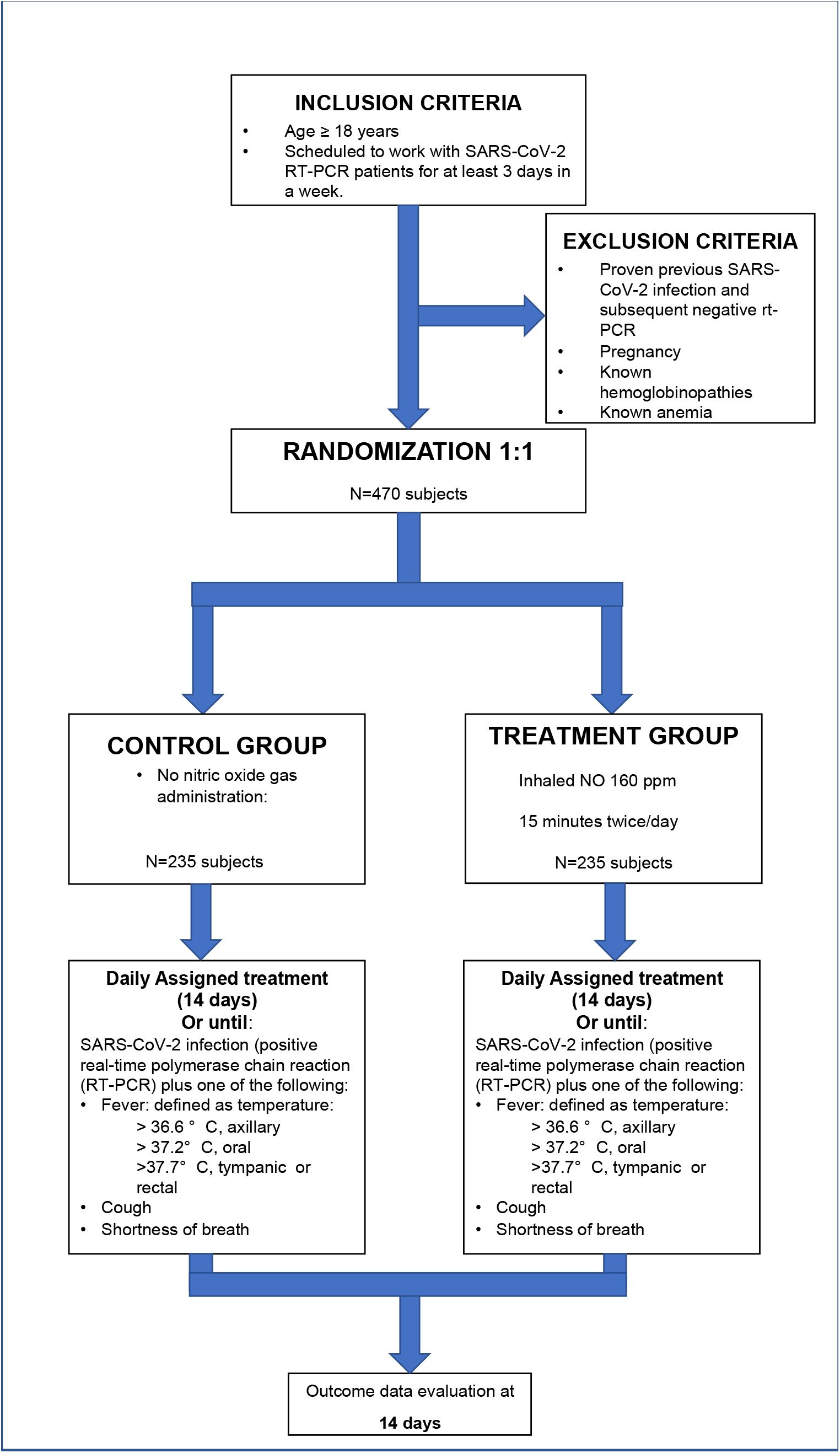
Study Flowchart

We will administer study gas through a mouthpiece connected to a breathing circuit (see Figure 2). This circuit is composed by an inspiratory (with NO, air and oxygen inlet) and an expiratory limb. On the inspiratory limb there are (I) a 3L reservoir bag and (II) a scavenger containing Soda Lime to minimize the NO_2_ concentration administered to the subject [13]. To avoid oxygen and NO mixing in the reservoir bag (leading to NO_2_ formation) a one-way valve is positioned between nitric oxide and oxygen inlet.

**Figure 2.**
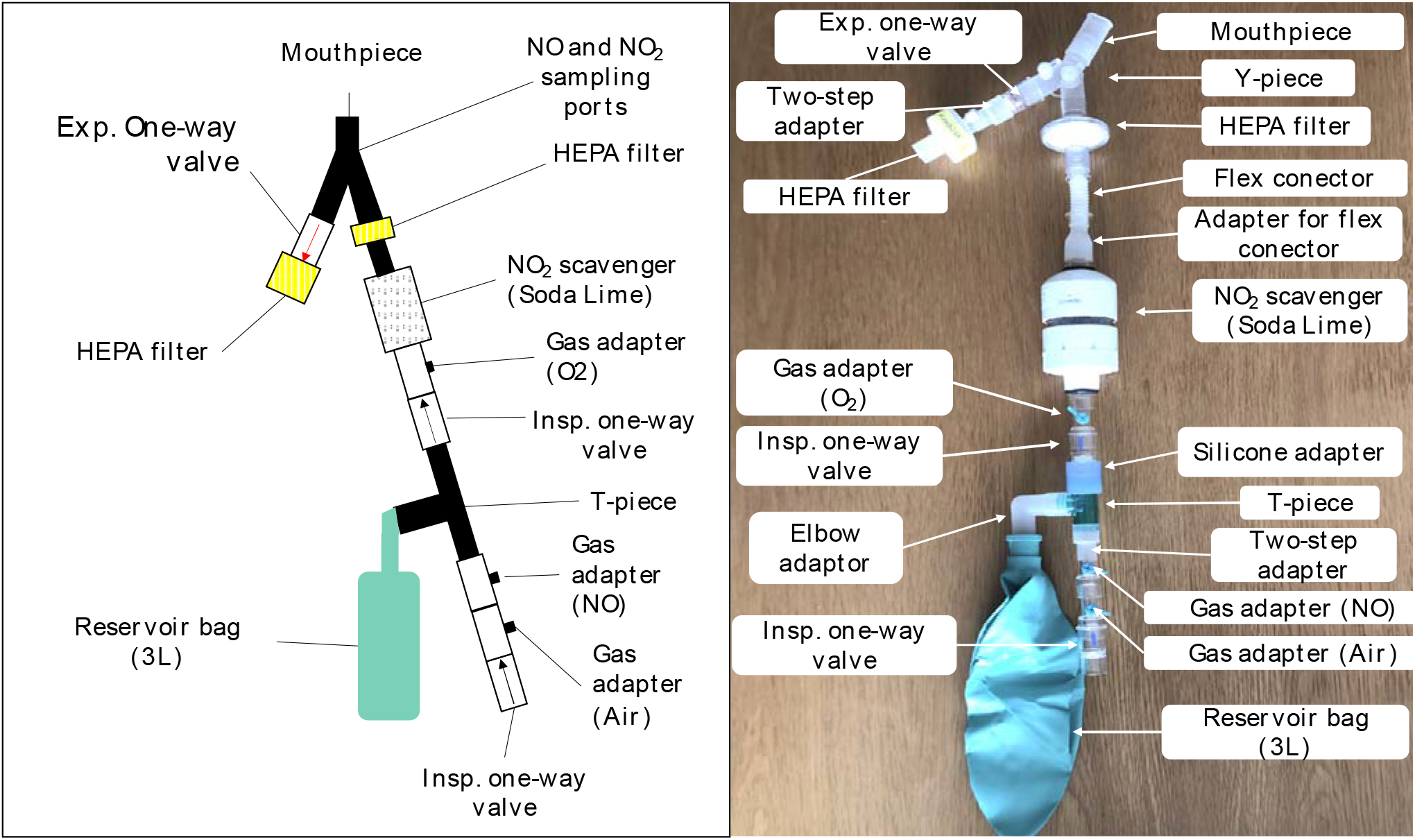
Nitric oxide gas delivery system.

Inspiratory concentration of NO and NO_2_ will be continuously monitored; NO_2_ concentrations will be maintained below 2 ppm. Methemoglobin will also be continuously monitored non- invasively with a dedicated pulse oximeter system and will be maintained below 5%. NO delivery will be titrated in order to keep methemoglobin under 5%.

During the session, SpO_2_ and heart rate will be continuously monitored by the study staff. Blood pressure will be measured non-invasively before and after the treatment. Values will be recorded in a dedicated paper data sheet, specifically: (I) before starting NO delivery, (II) at the end of the delivery, and (III) at 5 to 10 minutes after completion of the NO delivery to demonstrate a decrease of methemoglobin after cessation of gas delivery. Subjects assigned to the control group will not receive any gas therapy. Subjects will be monitored for 14 days in both groups.

### Safety

Breathing NO at 160 ppm for 30 minutes is safe [14]. The theoretical risks of NO breathing include the following: pulmonary edema, methemoglobinemia, hypoxia, and hypotension. In a small study of healthy volunteers, Frostell et al [15] reported that there were no adverse clinical events in the group inhaling NO. Of note, the average baseline methemoglobin level prior to NO administration was 0.61%. This increased to 0.77% after 10 minutes of inhalation of NO at 80 ppm. This was a statistically significant, although clinically irrelevant, increase in methemoglobin levels. In our recent study [16], there was no side effect or drug toxicity associated with the use of NO at 80 ppm for up to 24 hours.

During nitric oxide gas administration, the study staff will continuously monitor the NO, NO_2_, and oxygen concentrations in the inhaled gas. Peripheral oxygen saturation (SpO_2_) will be continuously monitored. Methemoglobin levels will be assessed using a non-invasive continuous co-oximetry monitor. If the methemoglobin level rises above 5%, NO will be progressively reduced to reach a value < 5%.

Any study subject who experiences a side effect that is suspected to be related to the study drug will not be allowed to continue in the protocol.

### Blinding

No masking is going to be used in this study. Subjects and investigators will be aware of the study allocation.

### Outcomes

The primary endpoint of this study is the incidence of subjects with COVID-19 disease 14 days post-enrollment. COVID-19 is defined as a SARS-CoV-2 infection confirmed by a positive real-time polymerase chain reaction (RT-PCR) with fever (> 36.6 C from the axillary site; or > 37.2 C from the oral site; or > 37.8 C from the rectal/tympanic site), cough, or shortness of breath.

The secondary endpoints are the proportion of healthcare providers who present a positive real time RT-PCR test for SARS-CoV-2 14 days after enrollment, proportion of healthcare providers requiring quarantine, and total number of quarantine days in the two groups.

### Sample size

Based on available data from Italy and China, we predict a 15% incidence of SARS-CoV-2 infection among healthcare providers and we assume that the incidence will be reduced to 5% with inhalant NO cycles. Considering an alpha level of 0.05 and a power of 0.9, we determined a sample size of n1=207, n2=207, by a two-sided test. In order to account for possible dropouts, we increased the sample size by about 10% from 414 to a total of 470 subjects (235 each group). Estimated sample sizes where calculated using the Stata 14.1 software.

### Recruitment

This study is targeted for a population of healthcare providers working in a hospital with COVID-19 patients. Upon admission of the first SARS-CoV-2 positive patient in a hospital unit, all the healthcare professionals that meet the inclusion and not the exclusion criteria become eligible for the study. The clinical staff of each unit will be adequately informed about the protocol details. No clinical care procedure will be interrupted or delayed due to the study procedure.

### Assignment of interventions

Randomization will occur through a random allocation sequence generated by a computerized random generation program (REDCap). Parallel allocation to the treatment or control groups will occur with a 1:1 ratio. Randomization will not be stratified for pre-specified demographic conditions (e.g., sex, age). Results will be adjusted for such conditions in the analysis phase as specified in the statistical analysis section.

Subjects will be randomly allocated to either the treatment group with NO administration or to the control group with no NO administration.

### Data collection methods

Clinical information including medical history will be obtained from the participant interview. Collection of study variables will be managed by the outcome assessors using a dedicated subject’s file on REDCap.

### Data management

Outcome assessors, treatment providers, and the principal investigator will obtain unique usernames and passwords to transfer all data to a REDCap database dedicated to the study.

### Statistical methods

Data will be analyzed following the intention to treat analysis principle. Demographic and clinical data will be presented as proportions for categorical outcomes and mean plus standard deviation or median plus interquartile range for continuous outcomes. Comparison between groups for the primary outcome will be made with the X^2^ test or the Fisher exact test for categorical variables. Secondary outcomes will be analyzed with the X^2^ test or the Fisher exact test for categorical variables and with the t-test or Wilcoxon rank-sum test for continuous variables after assessing the total number of quarantine days for normal distribution.

A subgroup analysis will be performed using multiple linear regression, as well as logistic and cox regression models to adjust for age, pulmonary comorbidities, history of neoplastic disease, and the hour of exposition to SARS-CoV-2 positive patients in 14 days.

### Data monitoring

Data will be monitored by the principal investigator (PI) in collaboration with an independent Data and Safety Monitoring Board (DSMB). The PI and DSMB will monitor adverse events, evaluate data quality, and provide recommendations accordingly. The PI will monitor compliance to safety rules every 20 patients. Every violation will be reported to the DSMB. Before the beginning of the study, the DSMB will meet to decide safety rules and stopping Guidelines.

The Data Safety Management Board (DSMB) will perform an interim analysis for superiority after the 25^th^ patient. In case of a significant decreased infection rate in the treated subjects, the trial will be stopped. The signed informed consent will be kept in a secure place for at least 5 years after study completion.

### Harm

Safety data includes levels of Methemoglobin, NO_2_ levels, minor deviation from the protocol, and minor adverse reactions to NO gas administration. Other data to be reviewed includes the maintenance of patient confidentiality throughout the study. In accordance with PHRC policy on Adverse Event Reporting and Unanticipated Problems Involving Risks to Subjects, the Principal Investigator will report adverse events or other unanticipated problems to the DSMB and to the PHRC within 5 working days/7 calendar days of the date the investigator first becomes aware of the problem. Mild or moderate adverse events will be presented in progress reports at continuing reviews.

The decision regarding altering or stopping the protocol will be performed by the principal investigator together with the Data Safety Management Board (DSMB). Protocol exit criteria will be:

a. Acute worsening hypotension defined by a decrease in mean blood pressure of > 20 mmHg not attributable to other causes such as the progression of the disease, hypovolemia, hemorrhage, sepsis, or acute heart failure.
b. Sudden hypoxemia defined as a 5% reduction of peripheral oxygen saturation (SpO_2_) from the basal value.
c. Any life-threatening symptom potentially attributed to NO administration by the physician investigator.

## Data Availability

Data sharing not applicable to this article as no datasets were generated or analysed during the current study.

## Ethics and dissemination

The study will be conducted following the Good Clinical Practices of the International Conference on Harmonization Good Clinical Practices (ICH-GCP) as well as the local and national regulations.The results of the study will be published in scientific journals and presented at scientific meetings. Personal information about enrolled participants will be collected only through the RedCap platform.

## Dissemination Policy

Results will be published and will be accessible to healthcare professionals and the public.

## Funding statement

Local departmental funds

## Competing interests’ statement

LB salaries are partially supported by NIH/NHLBI 1 K23 HL128882-01A1.

## Notes

### Competing Interest Statement

The authors have declared no competing interest.

### Clinical Trial

NCT04312243

## References

1. https://www.healthmap.org/covid-19/.

2. Wang, D., et al., Clinical Characteristics of 138 Hospitalized Patients With 2019 Novel Coronavirus-Infected Pneumonia in Wuhan, China. JAMA, 2020.

3. Zhou, P., et al., Protecting Chinese Healthcare Workers While Combating the 2019 Novel Coronavirus. Infection Control & Hospital Epidemiology, 2020: p. 1–4.

4. apollo.massgeneral.org/coronavirus

5. Privett, B.J., et al., Examination of bacterial resistance to exogenous nitric oxide. Nitric oxide : biology and chemistry, 2012. 26(3): p. 169–173.

6. McMullin, B.B., et al., The Antimicrobial Effect of Nitric Oxide on the Bacteria That Cause Nosocomial Pneumonia in Mechanically Ventilated Patients in the Intensive Care Unit. Respiratory Care, 2005. 50(11): p. 1451–1456.

7. Akerstrom, S., et al., Nitric oxide inhibits the replication cycle of severe acute respiratory syndrome coronavirus. J Virol, 2005. 79(3): p. 1966–9.

8. Akerstrom, S., et al., Dual effect of nitric oxide on SARS-CoV replication: viral RNA production and palmitoylation of the S protein are affected. Virology, 2009. 395(1): p. 1–9.

9. Keyaerts, E., et al., Inhibition of SARS-coronavirus infection in vitro by S-nitroso-N-acetylpenicillamine, a nitric oxide donor compound. Int J Infect Dis, 2004. 8(4): p. 223–6.

10. Chen, L., et al., Inhalation of nitric oxide in the treatment of severe acute respiratory syndrome: a rescue trial in Beijing. Clin Infect Dis, 2004. 39(10): p. 1531–5.

11. Deppisch, C., et al., Gaseous nitric oxide to treat antibiotic resistant bacterial and fungal lung infections in patients with cystic fibrosis: a phase I clinical study. Infection, 2016. 44(4): p. 513–20.

12. Regev-Shoshani, G., et al., Gaseous nitric oxide reduces influenza infectivity in vitro. Nitric Oxide, 2013. 31: p. 48–53.

13. Ishibe T, Sato T, Hayashi T, Kato N, Hata T. Absorption of nitrogen dioxide and nitric oxide by soda lime. Br J Anaesth. 1995 Sep;75(3):330–3.

14. Miller, C., et al., Gaseous nitric oxide bactericidal activity retained during intermittent high-dose short duration exposure. Nitric Oxide, 2009. 20(1): p. 16–23.

15. Frostell CG. et al. Inhaled nitric oxide selectively reverses human hypoxic pulmonary vasoconstriction without causing systemic vasodilation. Anesthesiology. 1993 Mar;78(3):427–435.

16. Lei C. et al. Nitric Oxide Decreases Acute Kidney Injury and Stage 3 Chronic Kidney Disease after Cardiac Surgery. Am J Respir Crit Care Med. 2018 Nov 15;198(10):1279–1287

